# Shared Inherited Genetics of Benign Prostatic Hyperplasia and Prostate Cancer

**DOI:** 10.1101/2021.12.10.21267604

**Authors:** Alexander Glaser, Zhuqing Shi, Jun Wei, Nadia A. Lanman, Skylar Ladson-Gary, Renee E Vickman, Omar E. Franco, Susan E. Crawford, S. Lilly Zheng, Simon W. Hayward, William B. Isaacs, Brian T. Helfand, Jianfeng Xu

**Affiliations:** Program for Personalized Cancer Care, NorthShore University HealthSystem, Evanston, IL; Department of Surgery, NorthShore University HealthSystem, Evanston, IL; Collaborative Core for Cancer Bioinformatics, Purdue Center for Cancer Research, Purdue University, West Lafayette, IN; Department of Urology and the James Buchanan Brady Urologic Institute, Johns Hopkins University School of Medicine; Baltimore, MD; Department of Surgery, University of Chicago Pritzker School of Medicine, Chicago, IL

**Keywords:** Benign prostatic hyperplasia (BPH), prostate cancer, heritability, genetic correlation, SNPs, genetic risk score, lethal

## Abstract

**Background:** The association between benign prostatic hyperplasia (BPH) and prostate cancer (PCa) remains controversial, largely due to inherent detection bias in traditional observational studies. The objective of this study is to assess their association using inherited SNPs.

**Methods:** Subjects were White men from the large population-based UK Biobank (UKB). Association between BPH and PCa was tested: 1) phenotypical correlation using chi-square test, 2) genetic correlation (*r*_g_) based on 1,126,841 polymorphic SNPs across the genome using linkage disequilibrium score regression (LDSR), and 3) cross-disease genetic associations based on known risk-associated SNPs (15 for BPH and 239 for PCa), individually and cumulatively as measured by genetic risk score (GRS).

**Findings:** Among 214,717 White men in the UKB, 24,623 (11.47%) and 14,311 (6.67%) had a diagnosis of BPH and PCa, respectively. Diagnoses of these two diseases were significantly correlated, *χ*^2^=1862.80, *P*<1E-299. A significant genetic correlation was found, *r*_g_ (95% confidence interval (CI))=0.27 (0.15-0.39), *P*=9.17E-06. In addition, significant cross-disease genetic associations for established risk-associated SNPs were also found. Among the 250 established GWAS-significant SNPs of PCa or BPH, 51 were significantly associated with risk of the other disease at *P*<0.05, significantly more than expected by chance (N=12), *P*=3.04E-7 (*χ*^2^-test). Furthermore, significant cross-disease GRS associations were also found; GRS_BPH_ was significantly associated with PCa risk (odds ratio (OR)=1.26 (1.18-1.36), *P*=1.62E-10), and GRS_PCa_ was significantly associated with BPH risk (OR=1.03 (1.02-1.04), *P*=8.57E-06). Moreover, GRS_BPH_ was significantly and inversely associated with lethal PCa risk in a PCa case-case analysis (OR=0.58 (0.41-0.81), *P*=1.57E-03). In contrast, GRS_PCa_ was not significantly associated with lethal PCa (OR=0.99 (0.94-1.04), *P*=0.79).

**Interpretation:** BPH and PCa share common inherited genetics which suggests the phenotypical association of these two diseases in observational studies is not entirely caused by detection bias. This novel finding may have implications in disease etiology and risk stratification.

**Funding:** None.

## Introduction

Benign prostatic hyperplasia (BPH) is a histological diagnosis characterized by a proliferation of both stromal and epithelial cells in the transitional zone of the prostate.^1^ This proliferation can lead to bladder outlet obstruction and subsequent lower urinary tract symptoms (LUTS). Prostate cancer (PCa), on the other hand, is a malignant adenocarcinoma primarily found in the peripheral zone of the prostate and, prior to metastasis is typically asymptomatic being primarily detected by PSA-screening.^2^ BPH and PCa are two of the most common diseases in men and incidence for both conditions increases considerably with age.^3, 4^ These two pathologic processes negatively impact quality of life and result in considerable healthcare expense.

Despite the major differences in cellular growth patterns and stereotyped locations within the prostate, a link between BPH and PCa has been hypothesized, studied and reported.^5, 6^ Their co-occurrence was first documented from autopsy studies in 1957 and 1992.^7, 8^ Since then, conflicting results on the association have been published in multiple epidemiological studies, including those from retrospective case-control studies,^9, 10^ prospective cohorts,^10^ secondary analysis of clinical trials,^11, 12^ large population-based cohorts,^13, 14^ and a recent meta-analysis.^15^ To date, no consensus has been reached on their association and causal relationship.^6^ Consequently, the current National Cancer Institute website states that ‘BPH is not linked to cancer and does not increase your risk of getting PCa’.^16^

A major cause for the inconclusive findings is the inherent detection bias of observational studies in diagnosing these two diseases. Patients treated by urologists for one of these diseases (e.g., BPH) are more likely to be examined thoroughly and are therefore more likely to be diagnosed for the other disease (PCa).^17^ This detection bias is particularly prominent because the likelihood of diagnosing BPH and PCa increases with heightened prostate examinations and diagnostic evaluations that include prostate-specific antigen (PSA) measurements, which is included in current guidelines for each of these disease states.^18, 19^

The availability of high-throughput SNP genotyping technology in the last 15 years enabled a systematic evaluation of genetic variants in the genome among large numbers of subjects. This technology advanced the understanding of the polygenic architecture of common diseases in two distinct ways. First, using millions of SNPs in the genome, it is now possible to estimate polygenic heritability of a disease using linkage disequilibrium score regression (LDSR).^20^ Furthermore, unbiased estimates of SNP-based genetic correlation between two diseases can be obtained using LDSR. Second, it is also feasible to systematically compare genetic variants in the genome in subjects with or without a disease using genome-wide association studies (GWAS) to identify disease risk-associated SNPs. Owing to the rigorous requirement of statistical significance (*P*<2E-08) and confirmation in independent studies, risk-associated SNPs identified by GWAS most likely represent true associations. To date, 269 and 17 risk-associated SNPs have been identified for PCa and BPH, respectively.^21, 22^ Although the effect of these risk-associated SNPs is moderate individually, they are additive and have a stronger cumulative effect that can be measured by a polygenic risk score. Published studies have consistently demonstrated the performance of polygenic risk score for measuring and stratifying inherited risk.^21-24^ Due to the inherited nature of SNPs (fixed at conception and always preceding occurrence of diseases), they are less susceptible to detection bias in assessing associations of two diseases from observational studies.

The primary hypothesis of this study is that BPH and PCa are linked and that this association is partially contributed by shared inherited genetics via the same genes (causal or pleotropic effects) and/or different genes in linkage disequilibrium (LD). This hypothesis was tested in a large population-based cohort by 1) assessing phenotypical associations between the two diseases, 2) estimating their genetic correlation based on millions of SNPs in the genome using LDSR, and 3) performing cross-disease genetic association tests based on known risk-associated SNPs, individually and cumulatively. Furthermore, the clinical utilities of polygenic risk score of BPH and PCa for predicting the risk of these two diseases were also assessed.

## Methods

### Subjects

Subjects of this study were from the UK Biobank (UKB), a population-based study with extensive genetic and phenotypic data for approximately 500,000 individuals from across the United Kingdom aged between 40 and 69 at recruitment (accessed under Application Number: 50295).^25^ Extensive phenotypic and genomic information is available for each participant in the UKB, including disease diagnosis, questionnaire, biological measurements, lifestyle indicators, and biomarkers in blood and urine. Diagnoses of BPH (Data-Field 132073) and the procedure for transurethral resection of the prostate (TURP) (Data-Field 41200, 41210 and 41272), as well as diagnosis of PCa (Data-Field 40001, 40002, 40006, 41202, 41204, 41270) were provided by the UKB based on the ICD-10 code and/or self-reports (released on July 9^th^, 2021). Information on PCa-specific death (lethal PCa) was based on death registries. Genome-wide SNPs data are available for all participants.

### SNPs and polygenic risk score

Independent risk-associated SNPs for BPH and PCa included in this study were established using evidence-based review of published GWAS (defined as *P*<5×10^-8^ and pair-wise linkage disequilibrium (LD, *r*^2^<0.2)) and are available in the UKB, including 15 for BPH and 239 for PCa. Their SNP ID, risk and reference alleles, odds ratio, allele frequency, and references are described in **sTable 1**.

The cumulative effect of SNPs on each disease was measured by genetic risk score (GRS), a population-standardized polygenic risk score. GRS was calculated by multiplying the per-allele odds ratio (OR) with number of risk alleles of each SNP and normalizing the risk by the average risk expected in the population.^26^ As such, GRS value can be interpreted as relative risk to the general population regardless number of risk-associated SNPs used in GRS calculation.

### Statistical analysis

Correlation between BPH and PCa was assessed using a chi-square test. The strength of association (OR and 95% confidence interval (CI)) between the two diseases was estimated using a multivariable logistic regression adjusting for age at recruitment and genetic background (top 10 principal components provided by the UKB).

SNP-based heritability (*h*^2^) for BPH and PCa, respectively, and genetic correlation (*r*_g_) between BPH and PCa were estimated based on polymorphic SNPs (minor allele frequency >0.01) in the genome using LDSR analysis.^20^ GWAS summary statistics of BPH and PCa (adjusting for age at recruitment and genetic background) were matched to the pre-computed LD scores of the 1000 Genomes European reference. SNP heritability estimates were converted to the liability-scale based on the observed prevalence in the UKB.

Cross-disease genetic association for BPH and PCa was tested based on known risk-associated SNPs (individually and cumulatively as measured by GRS) of these two diseases. Association of individual SNPs and GRS with cross-disease risk was tested adjusting for age at recruitment and genetic background.

## Results

Among 214,717 White men in the UKB, 24,623 (11.47%) and 14,311 (6.67%) had a diagnosis of BPH and PCa, respectively (**Table 1**). Diagnoses of these two diseases were significantly correlated, *χ*^2^=1862.80, *P*<1E-299. Specifically, 3,231 (1.50%) men had a diagnosis of both BPH and PCa, which was significantly higher than the 1,332 (0.62%) expected assuming that the diagnosis of these diseases were independent, *χ*^2^=797.96, *P*=1.50E-175. Having a diagnosis of PCa was associated with an OR (95% CI) of 1.58 (1.51-1.65) for BPH risk, *P*=5.13E-94. Conversely, having a diagnosis of BPH was associated with an OR (95%CI) of 1.57 (1.50-1.64) for PCa risk, *P*=1.07E-90. These ORs were estimated adjusting for age at recruitment and genetic background.

**Table 1.** Diagnosis of BPH and PCa among White men in the UKB (N=214,717)

|  |  | Median age (IQR), yr |  |
| --- | --- | --- | --- |
| Diagnosis | No. (%) of men | At recruitment | Age at diagnosis |
| BPH |  |  |  |
| Any BPH | 24,623 (11.47) | 63.5 (60.5-66.5) | 62.3 (57.31-67.19) |
| TURP* | 5,704 (26.61) | 64.5 (61.5-67.5) | 61.58 (57.07-66.31) |
| Non-TURP* | 15,833 (73.39) | 63.5 (59.5-66.5) | 62.57 (57.4-67.41) |
| PCa |  |  |  |
| Any PCa | 14,311 (6.67) | 63.5 (59.5-66.5) | 65.35 (60.08-69.16) |
| Lethal PCa | 692 (4.84) | 65.5 (62.5-68.5) | 65.5 (61.02-69.46) |
| Non-lethal PCa | 13,619 (95.16) | 63.5 (59.5-66.5) | 65.33 (60.8-69.12) |
| Both BPH and PCa | 3,231 (1.5) | 64.5 (61.5-67.5) | 62.57 (58.06-67.12)** |
| Neither BPH or PCa | 179,014 (83.37) | 57.5 (49.5-63.5) | \ |

\*Number of subjects with missing data for TURP (49,817)
\*\*Based on earlier age at diagnosis of BPH and PCa

Genetic susceptibility to BPH and PCa was estimated based on polymorphic SNPs across the genome (minor allele frequency >1%). SNP-based heritability (*h*^2^) (95% CI) was 0.09 (0.07-0.11) for BPH (*P*=0.005) and 0.15 (0.12-0.18) for PCa (*P*=4.87E-10) (**Figure 1**). Furthermore, a significant and positive genetic correlation between the two diseases was found, *r*_g_ (95% CI) was 0.27 (0.15-0.39), *P*=9.17E-06.

**Figure 1.**
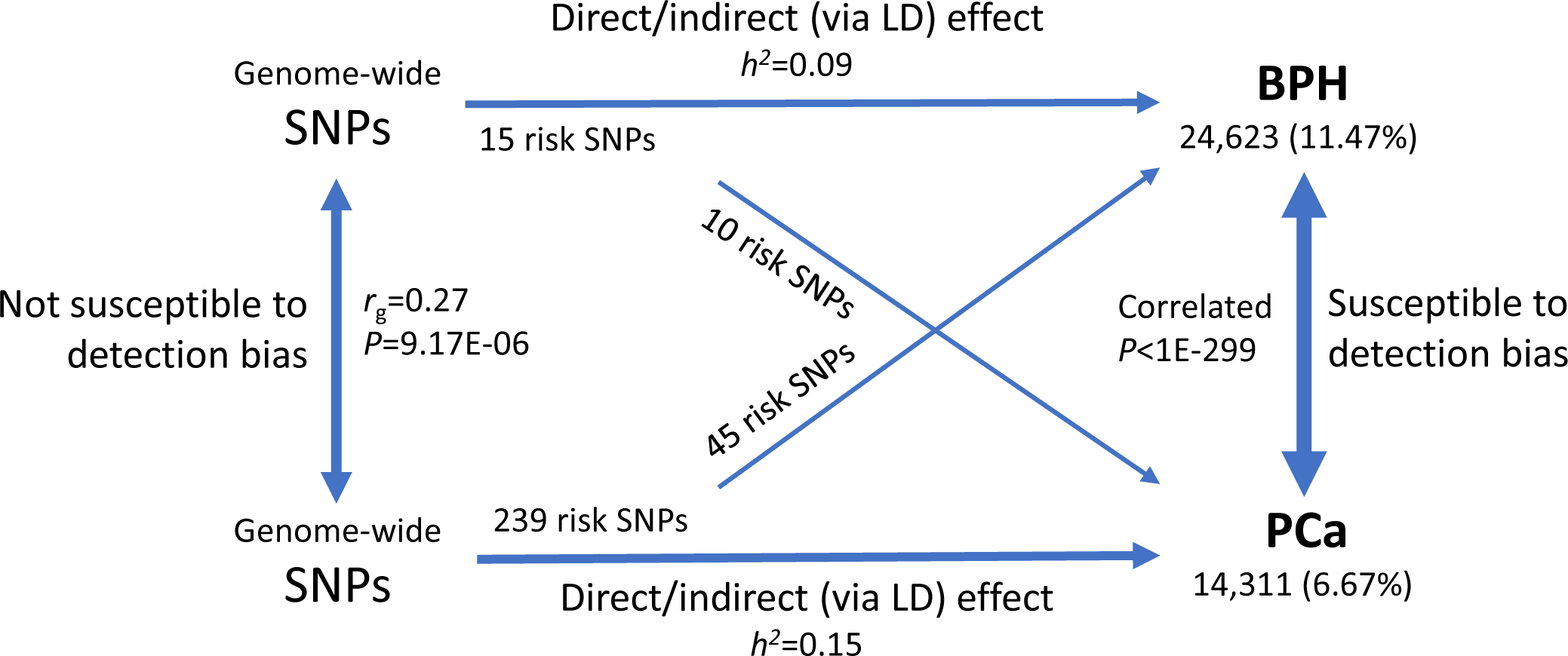
Summary of findings for the phenotypic and genetic link between BPH and PCa in White men of the UKB. First, diagnoses of BPH and PCa were significantly correlated (bi-direction arrows on the right side, *P*<1E-299). Second, inherited polygenic background contributed to the diagnosis of each disease (horizontal arrows, *h*^2^ of 0.09 and 0.16 for BPH and PCa, respectively). Third, polygenic background for BPH and PCa were significantly correlated (bi-direction arrows on the left side, *r*_g_=0.27, *P*=9.17E-06). Lastly, cross-disease association of established risk-associated SNPs for BPH and PCa (diagonal arrows). These findings provide strong statistical evidence that diagnoses of BPH and PCa are linked and the excessed co-occurrence of these two diseases was in part contributed by inherited genetics and not entirely driven by detection bias.

When examining established risk-associated SNPs for BPH and PCa, significant cross-disease genetic associations were found. Among the 250 risk-associated SNPs of PCa or BPH from previous GWAS studies, 51 were significantly associated with risk of the other disease at *P*<0.05, which was significantly more than expected by chance (N=12), *P*=3.04E-7 (*χ*^2^-test). Specifically, among the 239 established independent PCa risk-associated SNPs, 45 were associated with BPH diagnosis at *P*<0.05 (**sTable 1**). Reciprocally, among the 15 established independent BPH risk-associated SNPs, 10 were associated with PCa diagnosis at *P*<0.05. It is noted that 4 established GWAS-significant risk-associated SNPs of PCa and BPH were overlapped.

We also evaluated the cumulative effect of SNPs on disease risk. In addition to highly significant association between disease-specific GRS and their respective disease risks, significant cross-disease associations were also found (**Table 2**). GRS based on the 15 established BPH risk-associated SNPs was positively associated with PCa diagnosis (OR=1.26, *P*=1.62-E-10). Similarly, GRS based on the 239 established PCa risk-associated SNPs (GRS_PCa_) was significantly associated with BPH diagnosis (OR=1.03, *P*=8.57E-06).

**Table 2.** Performance of GRS for predicting disease risk in the UK Biobank, N=214,717

| GRS (No. risk SNPs) | Comparison | Sample size | Association test* |  |
| --- | --- | --- | --- | --- |
|  |  |  | OR (95% CI) | P |
| BPH (15 SNPs) | BPH vs. controls | 24,623 vs 190,094 | 2.39 (2.26-2.52) | 5.23E-211 |
|  | BPH-TURP vs. controls | 5,704 vs 205,927 | 3.61 (3.26-3.98) | 3.76E-140 |
|  | BPH-TURP vs. BPH non-TURP | 5,704 vs 15,833 | 2.01 (1.78-2.26) | 9.65E-31 |
|  | PCa vs. controls | 14,311 vs 200,406 | 1.26 (1.18-1.36) | 1.62E-10 |
|  | Lethal PCa vs. controls | 692 vs 214,025 | 0.74 (0.53-1.02) | 0.07 |
|  | Lethal PCa vs. non-lethal PCa | 692 vs 13,619 | 0.58 (0.41-0.81) | 1.57E-03 |
| PCa (239 SNPs) | BPH vs. controls | 24,623 vs 190,094 | 1.03 (1.02-1.04) | 8.57E-06 |
|  | BPH-TURP vs. controls | 5,704 vs 205,927 | 1.04 (1.02-1.07) | 1.02E-03 |
|  | BPH-TURP vs. BPH non-TURP | 5,704 vs 15,833 | 1.02 (0.99-1.06) | 0.13 |
|  | PCa vs. controls | 14,311 vs 200,406 | 1.53 (1.51-1.55) | <1E-299 |
|  | Lethal PCa vs. controls | 692 vs 214,025 | 1.25 (1.21-1.29) | 2.74E-34 |
|  | Lethal PCa vs. non-lethal PCa | 692 vs 13,619 | 0.99 (0.94-1.04) | 0.79 |

\*Adjusting for age at recruitment and genetic background (top 10 principal components)

Furthermore, GRS was also associated with specific phenotypes of BPH and PCa (**Table 2**). Notably, GRS_BPH_ was inversely associated with lethal PCa risk when comparing lethal PCa cases vs. non-lethal PCa cases (OR=0.58, *P*=1.57E-03). Upon examining results by GRS_BPH_ deciles (**sTable 2**), while no trend of lethal PCa prevalence with GRS_BPH_ deciles was found (*P*=0.14), the prevalence of lethal PCa in men of the highest GRS_BPH_ decile was 0.23%, noticeably lower than that of remaining deciles (0.33%, *P*=0.02). In contrast, the prevalence of non-lethal PCa increased slightly with higher GRS_BPH_ deciles (*P*_trend_=0.01), with the highest rate (6.64%) found in men of the top GRS_BPH_ decile. As for GRS_PCa_, no association with lethal PCa was found in a case-case analysis (OR=0.99, *P*=0.79) GRS_PCa_, it was not associated with lethal PCa in case-case analysis (OR=0.99, *P*=0.79). The prevalence of both lethal PCa and non-lethal PCa increased by each decile at a similar scale (**sTable 2**).

In light of the genetic correlation and cross-disease association of GRS between these two diseases, we next explored the clinical utility of using two GRS values for predicting diagnosis of BPH, PCa, and both diseases (**Table 3, Figure 2**). For example, using a GRS of 1.5 as a cutoff value (i.e., 1.5-fold increased risk over the general population), men with both high GRS_BPH_ and GRS_PCa_ had a considerably higher prevalence of these two diseases compared to men with both low GRS_BPH_ and GRS_PCa_; 15.64% vs. 5.12% for PCa, 16.87% vs. 11.18% for BPH, and 3.76% vs. 1.19% for both BPH and PCa. Similarly, men with either high GRS_BPH_ or GRS_PCa_ had higher prevalence of these two diseases. For lethal PCa, men with low GRS_BPH_ and high GRS_PCa_ had a higher prevalence (0.68%) than those with both low GRS (0.25%) and especially those with high GRS_BPH_ but low GRS_PCa_ (0.11%). Regarding surgical intervention for BPH (TURP), men with high GRS_BPH_ had a higher prevalence of TURP than those with low GRS_BPH_, regardless of GRS_PCa_ (6.54% and 7.40% for high GRS_BPH_ vs. 3.29% and 3.57% for low GRS_BPH_).

**Figure 2.**
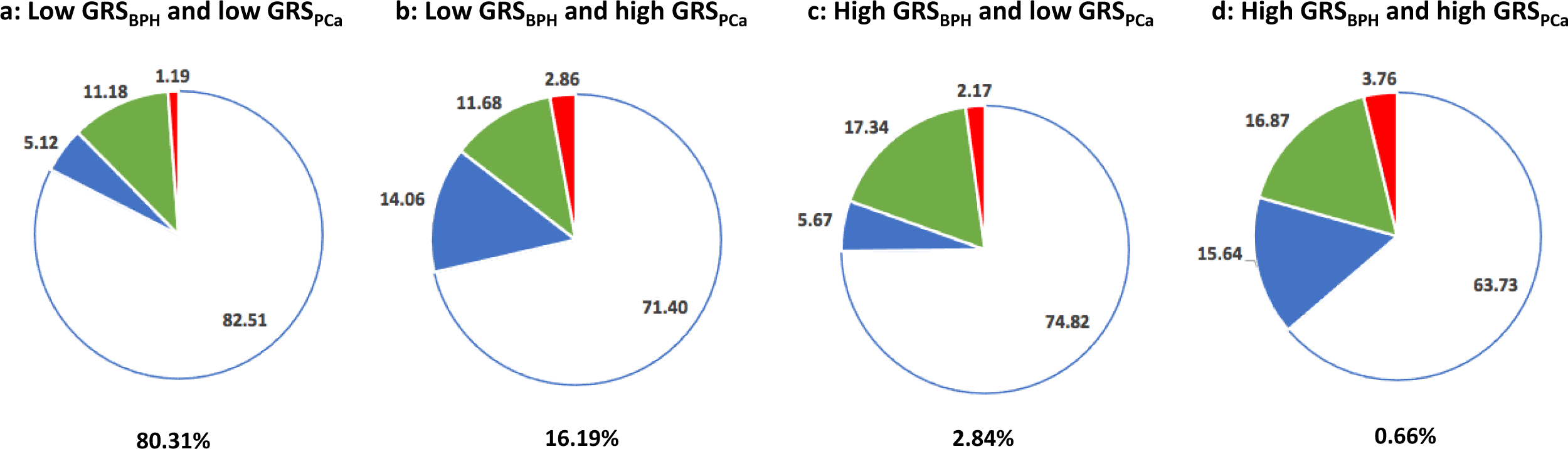
Pie charts of prevalence for BPH and PCa diagnoses in four groups of men with low (<1.5) and high (≥1.5) GRS_BPH_ and GRS_PCa_: a) both low, b) low GRS_BPH_ and high GRS_PCa_, c) high GRS_BPH_ and low GRS_PCa_, and d) both high. The percentage of men is each GRS group is indicated under each pie chart. Blue, green, and red slices represent the prevalence of PCa, BPH, and both diseases, respectively.

## Discussion

The primary goal of this study is to address the longstanding controversy surrounding the link between BPH and PCa.^5-15^ This clinically important question is complicated primarily by the inherent detection bias in epidemiological studies where patients diagnosed with one disease are typically examined more thoroughly by urologists and therefore have a higher chance of being diagnosed with the other disease.^17^ It is practically impossible to estimate the degree (partially or total) of the detection bias contributing to the observed association of BPH and PCa in traditional studies. Utilizing a large population-based cohort of over 200,000 men with diagnostic information for both diseases and genome-wide SNP data, we are able to test the association using both traditional epidemiological and alternative inherited genetic approaches (**Figure 1**). We 1) demonstrated a statistical association of phenotypical diagnoses of BPH and PCa (*P*<1E-299), 2) revealed a polygenic inherited basis for each of these two diseases (*h*^2^ of 0.09 and 0.16 for BPH and PCa, respectively), and 3) more importantly, the present analysis suggests that these two diseases are genetically correlated and that they share part of a polygenic background (*r*_g_=0.27) and risk-associated SNPs. A genetic correlation measures the proportion of the heritability (*h*^2^) that is shared between two diseases divided by the square root of the product of the heritability for each disease. This alternative inherited genetic approach does not directly test phenotypical co-occurrence of two diseases. Instead, it tests the correlation between heritability of each individual disease which is estimated only from the respective diseases, and therefore is not susceptible to detection bias. These findings provide strong statistical evidence that diagnoses of BPH and PCa are linked and the excessed concurrence of these two diseases was in part contributed by inherited genetics and not entirely driven by detection bias.

Shared inherited risk between BPH and PCa may arise from several potential sources, including 1) LD of different genes for BPH and PCa in the same chromosomal region, 2) pleiotropy where a single gene or variant affects both BPH and PCa, 3) causal effect where a gene causes a disease first which in turn causes the other disease, and 4) biases from population stratification. While the likelihood for the last source is low because this study was based on White men from a population-based cohort and the analysis adjusted for population stratification, we cannot differentiate among the sources of LD, pleiotropy and causality. Therefore, we can only conclude that shared inherited risk is directly or indirectly associated with both BPH and PCa but does not necessary directly cause these two diseases.

Although Mendelian randomization (MR) analysis is a well-established method utilizing measured inherited germline variations (genetic instruments) to interrogate the causal effect of an exposure on an outcome,^27^ we did not perform an MR analysis in this study for the following two considerations. First, considering the major difference between the two diseases in pathophysiology (benign proliferation for BPH and malignant growth for PCa), anatomic locations (primarily transition zone for BPH and peripheral zone for PCa), and cell types (epithelial and stromal hyperplasia for BPH; epithelial malignant transformation for PCa), we do not hypothesize that the development of either one of these two diseases is an exposure that causes the other disease. Second, it is challenging to test the validity of key assumptions underlying MR analysis (independent and exclusion).^28^ The independent assumption states that there are no unmeasured confounders of the associations between genetic variants and outcome. The exclusion restriction assumption requires that the genetic variants affect the outcome only through their effect on the risk factor of interest. Therefore, it is difficult to interpret MR results even if they are statistically significant.

As a comparison, we performed a similar analysis to assess the association and genetic correlation between BPH and another urological cancer (bladder cancer). While the diagnoses of these two diseases were highly correlated (*χ*^2^=1615.00, *P*<1E-299), there was no significant genetic correlation between the two diseases (*r*_g_=0.01, *P*=0.93). These results suggest that unlike BPH and PCa where the observed link is partially explained by shared inherited risk, there is no evidence that BPH and bladder share inherited genetic risk. The observed link between BPH and bladder cancer may be largely or completely caused by detection bias.

The finding of shared inherited risk between BPH and PCa from this study may have implications in understanding etiology and clinical utility. For example, identifying commonality of genes in the chromosomal regions that are associated with both BPH and PCa may help to better understand etiology for these two diseases. Toward this effort, we performed a preliminary pathway analysis for the 65 nearest genes in the 51 independent regions associated with both BPH and PCa using KEGG (Kyoto Encyclopedia of Genes and Genomes) (**sTable 3**). These genes are significantly enriched in only one biological pathway, the PCa pathway, *P*=0.007 (Benjamini correction) (**sTable 4**). Five (*AR, KLK3, TP53, FGFR2*, and *NKX3-1*) of the 65 genes (7.69%) are involved in the PCa pathway, significantly more than expected by chance (88 of 6,879 genes (1.28%) in the KEGG are in the PCa pathway). Similar results were found in the Ingenuity Pathway Analysis (version 01-20-04, Qiagen, CA). A major caveat of this analysis is that nearest genes may or may not account for the observed genetic associations, a common challenge for understanding biological mechanisms of GWAS findings.

The genetic correlation between the two diseases, especially cross-disease genetic association, also suggest GRS_BPH_ and GRS_PCa_ may be used in the clinic to stratify risk for these two diseases. For example, men with either or both high GRS_BPH_ and GRS_PCa_ have higher risks for these two diseases, alone or both, and men with high GRS_BPH_ are more likely to undergo surgical intervention. Men identified as high risk for these disease states may benefit from more aggressive screening and/or earlier referral to urologic sub-specialists.

While confirming the previous null result that GRS_PCa_ does not differentiate aggressiveness of the disease in PCa patients,^21, 24, 29^ we obtained a novel finding that GRS_BPH_ is inversely associated with lethal PCa in a case-case analysis. Specifically, men with the highest GRS_BPH_ decile have lower risk for lethal PCa but similar risk for non-lethal PCa when comparing to men in the remaining deciles. This finding is important and plausible considering that 1) GRS_BPH_ is positively associated with prostate volume,^22^ and 2) prostate volume is inversely associated with aggressive PCa.^30^ However, additional studies are needed to confirm this novel finding before exploring its clinical utility. Furthermore, more studies are warranted to understand specific SNPs and genes underlying this association and their biological effects on lethal PCa. Large PCa patient cohorts with germline data, PSA measurement, prostate volume, Gleason grade at time of diagnosis and prostatectomy surgery, as well as long-term disease follow-up are crucial to confirm and understand the association.

Several limitations of this study are noted. First, due to the fact that ∼95% of subjects in the UKB are White, our analyses and conclusions were limited to this group only. The generalizability of our finding in other ethnic and racial populations needs to be evaluated. Second, it is recognized that the UKB cohort was included in a previous GWAS that identified BPH risk-associated SNPs.^22^ Although this may inflate performance of GRS_BPH_ for predicting BPH risk, it has limited impact on the key findings of genetic correlation. Third, a lack of detailed clinical variables relevant to BPH and PCa in this population-based biobank such as PSA measurement, prostate volume, Gleason grade, and quantification of lower urinary tract symptoms hinder more granular analysis and understanding of clinical and genetic associations. Some findings from this study, especially the inverse association between GRS_BPH_ and lethal PCa, should be considered as preliminary. Comprehensive analysis in large and clinically well-characterized urological patient cohorts are needed. Finally, we did not perform functional analysis to investigate the biological mechanism of the genetic associations for both BPH and PCa. This is in part due to the nature of this genetic study where the primary goal is to identify chromosomal regions with statistical evidence for association. More importantly, we recognize the substantial challenges in these functional studies. The chromosomal region for each genetic association is typically large (>500 kb) and many of these regions are outside of coding genes. Nevertheless, we feel genetic association studies provide critical data relevant to human diseases. Geneticists and biologists can collaborate to understand biological mechanisms of genetic associations.

## Conclusions

In conclusion, utilizing genome-wide SNP data from a large population-based cohort demonstrated that BPH and PCa share common polygenic inherited risk. This novel genetic result suggests the excess of co-occurrence of these two diseases is not completely driven by detection bias. The current conclusion that BPH and PCa are not related, as stated by the NCI and other authoritative agencies, may be reconsidered.

## Data Availability

The data used in this study is available in the UK Biobank, a publicly available repository. Data was accessed through a Material Transfer Agreement under Application Reference Number: 50295. For additional information, please feel free to contact the corresponding author, Jianfeng Xu, DrPH.

## Acknowledgements

We are grateful to the Ellrodt-Schweighauser family for establishing Endowed Chair of Cancer Genomic Research (Xu), and Chez and Melman families for establishing Endowed Chairs of Personalized Prostate Cancer Care (Helfand), as well as the Rob Brooks Fund for Personalized Prostate Cancer Care at NorthShore University HealthSystem.

## Author Contributions

***Concept and design***: Xu, Helfand, Glaser

***Data analysis***: Shi, Wei, Lanman

***Manuscript draft***: Xu, Helfand, Glaser

***Critical revision of the manuscript for important intellectual content***: Glaser, Shi, Wei, Lanman, Ladson-Gary, Vickman, Franco, Crawford, Zheng, Hayward, Isaacs, Helfand, Xu

***Supervision***: Xu and Helfand

## Ethical approval and consent to participate

The UK Biobank was approved by North West – Haydock Research Ethics Committee (REC reference: 16/NW/0274; IRAS project ID: 200778). Data from the UK Biobank was accessed through a Material Transfer Agreement under Application Reference Number: 50295. This study was performed in accordance with the Declaration of Helsinki.

---

**sTable 1.**
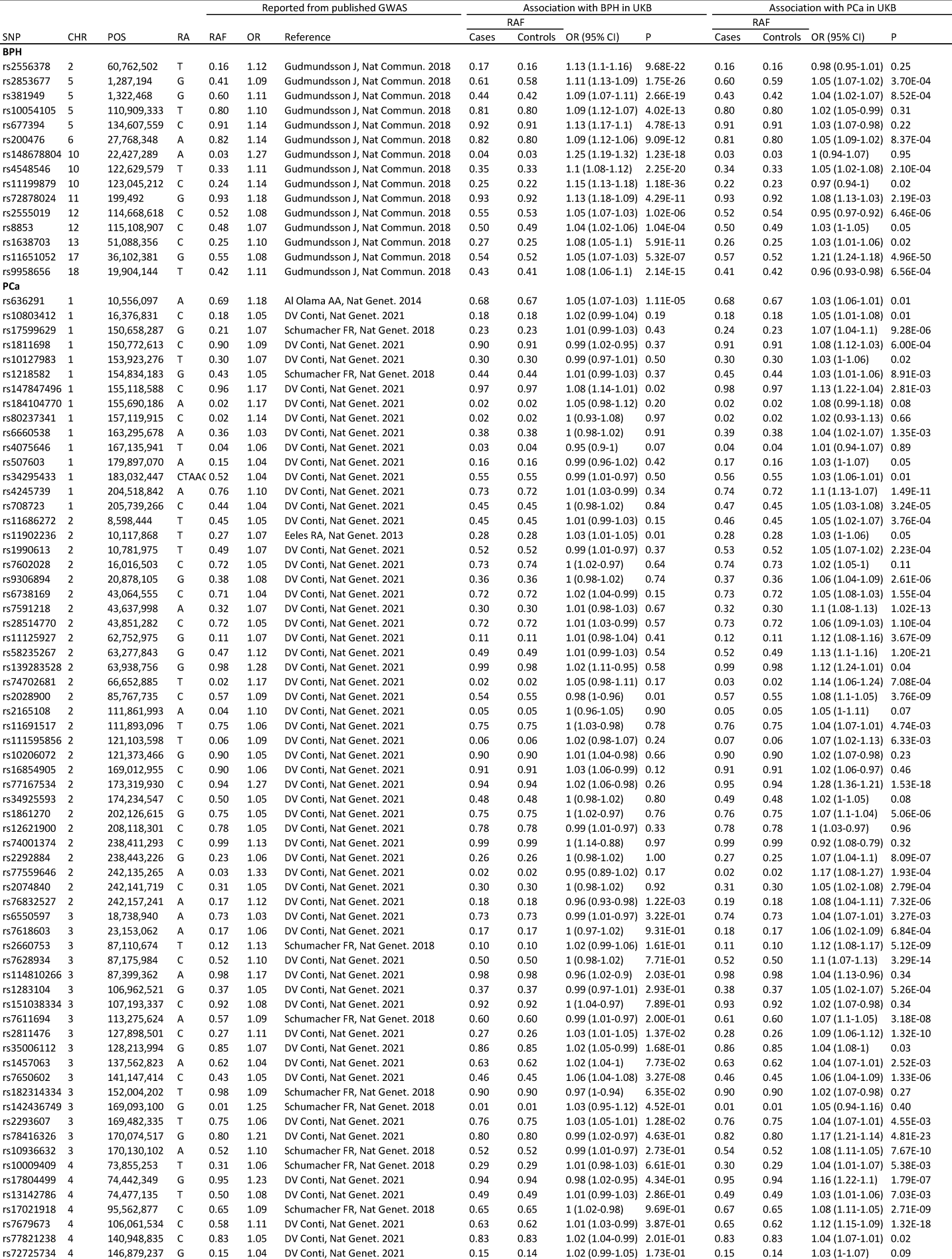
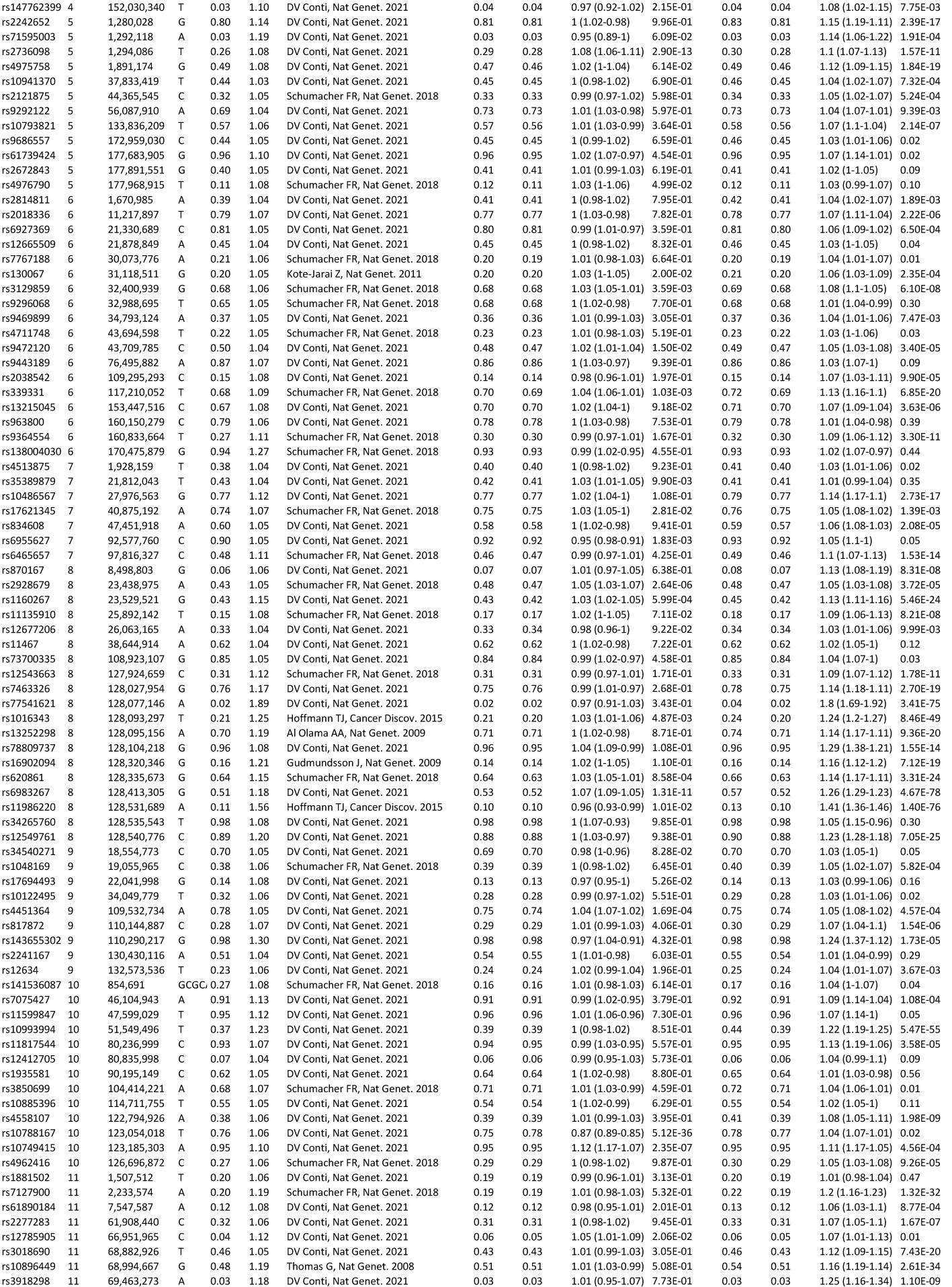
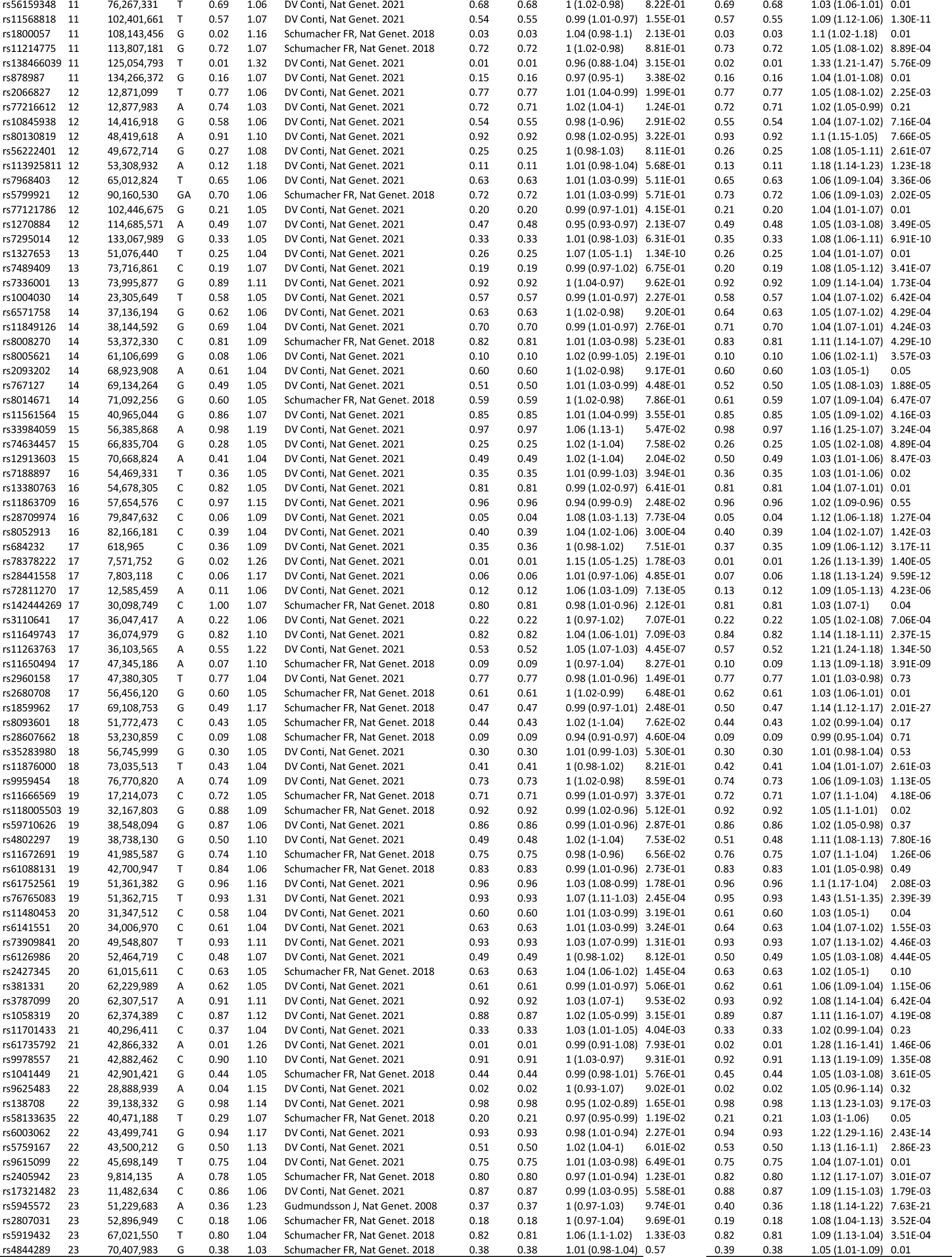
Established risk-associated SNPs for BPH and Pca

**sTable 2.** Prevalence of lethal and non-lethal PCa by GRS deciles

| GRS (range) | No. | Lethal PCa |  |  | Non-lethal PCa |  |  |
| --- | --- | --- | --- | --- | --- | --- | --- |
|  |  | No. | % | P-trend | No. | % | P-trend |
| GRS<sub>BPH</sub> |  |  |  | 0.14 |  |  | 0.01 |
| (0,0.704] | 19,783 | 61 | 0.31% |  | 1,183 | 5.98% |  |
| (0.704,0.783] | 19,780 | 62 | 0.31% |  | 1,190 | 6.02% |  |
| (0.783,0.845] | 19,792 | 81 | 0.41% |  | 1,248 | 6.31% |  |
| (0.845,0.902] | 19,784 | 62 | 0.31% |  | 1,265 | 6.39% |  |
| (0.902,0.96] | 19,770 | 66 | 0.33% |  | 1,267 | 6.41% |  |
| (0.96,1.02] | 19,782 | 59 | 0.30% |  | 1,283 | 6.49% |  |
| (1.02,1.09] | 19,779 | 67 | 0.34% |  | 1,241 | 6.27% |  |
| (1.09,1.18] | 19,786 | 64 | 0.32% |  | 1,305 | 6.60% |  |
| (1.18,1.31] | 19,779 | 63 | 0.32% |  | 1,210 | 6.12% |  |
| (1.31,3] | 19,775 | 45 | 0.23% |  | 1,313 | 6.64% |  |
| GRS<sub>PCa</sub> |  |  |  | 7.64E-47 |  |  | <1E-299 |
| (0,0.242] | 20,939 | 17 | 0.08% |  | 330 | 1.58% |  |
| (0.242,0.343] | 20,939 | 28 | 0.13% |  | 605 | 2.89% |  |
| (0.343,0.443] | 20,938 | 41 | 0.20% |  | 737 | 3.52% |  |
| (0.443,0.551] | 20,939 | 44 | 0.21% |  | 867 | 4.14% |  |
| (0.551,0.677] | 20,938 | 64 | 0.31% |  | 1,026 | 4.90% |  |
| (0.677,0.831] | 20,939 | 49 | 0.23% |  | 1,203 | 5.75% |  |
| (0.831,1.04] | 20,938 | 75 | 0.36% |  | 1,429 | 6.82% |  |
| (1.04,1.36] | 20,939 | 80 | 0.38% |  | 1,707 | 8.15% |  |
| (1.36,1.97] | 20,938 | 108 | 0.52% |  | 2,124 | 10.14% |  |
| (1.97,51] | 20,939 | 161 | 0.77% |  | 3,259 | 15.56% |  |

**sTable 3.** Nearest genes in the chromosomal regions that are associated with both BPH and PCa risk

| rsID | CHR | Position | Nearest genes | Gene Name |
| --- | --- | --- | --- | --- |
| rs636291 | 1 | 10,556,097 | PEX14 | peroxisomal biogenesis factor 14(PEX14) |
| rs147847496 | 1 | 155,118,588 | DPM3 | dolichyl-phosphate mannosyltransferase subunit 3(DPM3) |
| rs11902236 | 2 | 10,117,868 | GRHL1 | grainyhead like transcription factor 1(GRHL1) |
| rs2028900 | 2 | 85,767,735 | MAT2A | methionine adenosyltransferase 2A(MAT2A) |
| rs76832527 | 2 | 242,157,241 | ANO7 | anoctamin 7(ANO7) |
| rs2811476 | 3 | 127,898,501 | EEFSEC | eukaryotic elongation factor, selenocysteine-tRNA specific(EEFSEC) |
| rs7650602 | 3 | 141,147,414 | ZBTB38 | zinc finger and BTB domain containing 38(ZBTB38) |
| rs2293607 | 3 | 169,482,335 | TERC | telomerase RNA component(TERC) |
| rs2736098 | 5 | 1,294,086 | TERT | telomerase reverse transcriptase(TERT) |
| rs381949 | 5 | 1,322,468 | CLPTM1L | CLPTM1 like(CLPTM1L) |
| rs4976790 | 5 | 177,968,915 | COL23A1 | collagen type XXIII alpha 1 chain(COL23A1) |
| rs200476 | 6 | 27,768,348 | HIST1H2BL<br>LOC100131289 | histone cluster 1 H2B family member I(HIST1H2BL)<br>uncharacterized LOC100131289(LOC100131289) |
| rs130067 | 6 | 31,118,511 | CCHCR1 | coiled-coil alpha-helical rod protein 1(CCHCR1) |
| rs3129859 | 6 | 32,400,939 | BTNL2 | butyrophilin like 2(BTNL2) |
|  |  |  | HLA-DRA | major histocompatibility complex, class II, DR alpha(HLA-DRA) |
| rs9472120 | 6 | 43,709,785 | MRPS18A | mitochondrial ribosomal protein S18A(MRPS18A) |
|  |  |  | VEGFA | vascular endothelial growth factor A(VEGFA) |
| rs339331 | 6 | 117,210,052 | RFX6 | regulatory factor X6(RFX6) |
| rs35389879 | 7 | 21,812,043 | DNAH11 | dynein axonemal heavy chain 11(DNAH11) |
| rs17621345 | 7 | 40,875,192 | SUGCT | succinyl-CoA:glutarate-CoA transferase(SUGCT) |
| rs6955627 | 7 | 92,577,760 | LOC101927497<br>SAMD9 | uncharacterized LOC101927497(LOC101927497)<br>sterile alpha motif domain containing 9(SAMD9) |
| rs2928679 | 8 | 23,438,975 | NKX3-1<br>SLC25A37 | NK3 homeobox 1(NKX3-1)<br>solute carrier family 25 member 37(SLC25A37) |
| rs1016343 | 8 | 128,093,297 | PRNCR1 | prostate cancer associated non-coding RNA 1(PRNCR1) |
| rs620861 | 8 | 128,335,673 | CASC21 | cancer susceptibility candidate 21 (non-protein coding)(CASC21) |
| rs6983267 | 8 | 128,413,305 | CCAT2 | colon cancer associated transcript 2 (non-protein coding)(CCAT2) |
| rs11986220 | 8 | 128,531,689 | CASC11<br>CASC8 | cancer susceptibility candidate 11 (non-protein coding)(CASC11)<br>cancer susceptibility candidate 8 (non-protein coding)(CASC8) |
| rs4451364 | 9 | 109,532,734 | LINC01505<br>ZNF462 | long intergenic non-protein coding RNA 1505(LINC01505)<br>zinc finger protein 462(ZNF462) |
| rs10788167 | 10 | 123,054,018 | FGFR2<br>MIR5694 | fibroblast growth factor receptor 2(FGFR2)<br>microRNA 5694(MIR5694) |
| rs72878024 | 11 | 199,492 | ODF3 | outer dense fiber of sperm tails 3(ODF3) |
| rs12785905 | 11 | 66,951,965 | KDM2A | lysine demethylase 2A(KDM2A) |
| rs878987 | 11 | 134,266,372 | B3GAT1 | beta-1,3-glucuronyltransferase 1(B3GAT1) |
| rs10845938 | 12 | 14,416,918 | ATF7IP<br>GRIN2B | activating transcription factor 7 interacting protein(ATF7IP)<br>glutamate ionotropic receptor NMDA type subunit 2B(GRIN2B) |
| rs1270884 | 12 | 114,685,571 | RBM19<br>TBX5 | RNA binding motif protein 19(RBM19)<br>T-box 5(TBX5) |
| rs8853 | 12 | 115,108,907 | TBX3 | T-box 3(TBX3) |
| rs1327653 | 13 | 51,076,440 | DLEU1-AS1<br>ST13P4 | DLEU1 antisense RNA 1(DLEU1-AS1)<br>suppression of tumorigenicity 13 (colon carcinoma) (Hsp70 interacting protein) pseudogene 4(ST13P4) |
| rs12913603 | 15 | 70,668,824 | SALRNA3<br>TLE3 | senescence associated long non-coding RNA 3(SALRNA3)<br>transducin like enhancer of split 3(TLE3) |
| rs11863709 | 16 | 57,654,576 | ADGRG1 | adhesion G protein-coupled receptor G1(ADGRG1) |
| rs28709974 | 16 | 79,847,632 | LINC01229<br>LOC102724084 | long intergenic non-protein coding RNA 1229(LINC01229)<br>uncharacterized LOC102724084(LOC102724084) |
| rs8052913 | 16 | 82,166,181 | HSD17B2<br>MPHOSPH6 | hydroxysteroid 17-beta dehydrogenase 2(HSD17B2)<br>M-phase phosphoprotein 6(MPHOSPH6) |
| rs78378222 | 17 | 7,571,752 | TP53 | tumor protein p53(TP53) |
| rs72811270 | 17 | 12,585,459 | LOC101928418 | uncharacterized LOC101928418(LOC101928418) |
| rs11649743 | 17 | 36,074,979 | HNF1B | HNF1 homeobox B(HNF1B) |
| rs9958656 | 18 | 19,904,144 | CTAGE1<br>GATA6 | cutaneous T-cell lymphoma-associated antigen 1(CTAGE1)<br>GATA binding protein 6(GATA6) |
| rs28607662 | 18 | 53,230,859 | TCF4 | transcription factor 4(TCF4) |
| rs76765083 | 19 | 51,362,715 | KLK3 | kallikrein related peptidase 3(KLK3) |
| rs2427345 | 20 | 61,015,611 | GATA5<br>RBBP8NL | GATA binding protein 5(GATA5)<br>RBBP8 N-terminal like(RBBP8NL) |
| rs11701433 | 21 | 40,296,411 | LOC400867 | uncharacterized LOC400867(LOC400867) |
| rs58133635 | 22 | 40,471,188 | LOC100130899<br>TNRC6B | uncharacterized LOC100130899(LOC100130899)<br>trinucleotide repeat containing 6B(TNRC6B) |
| rs5919432 | 23 | 67,021,550 | AR<br>OPHN1 | androgen receptor(AR)<br>oligophrenin 1(OPHN1) |

**sTable 4.** Enrichment analysis of nearest genes in the chromosomal regions associated with both BPH and PCa (6,879 genes in KEGG)

| KEGG pathway (No. genes in the pathway) | No. (%) genes observed | Observed genes | P-Fisher | P-Benjamini |
| --- | --- | --- | --- | --- |
| Prostate cancer (88) | 5 (7.7%) | *AR, KLK3, TP53, FGFR2, NKX3-1* | 4.10E-06 | 7.10E-03 |
| Pathways in cancer (393) | 6 (9.2%) | *AR, KLK3, TP53, FGFR2, NKX3-1, VEGFA* | 6.50E-04 | 0.15 |
| Systemic lupus erythematosus (134) | 3 (4.6%) | *HIST1H2BL, HLA-DRA, GRIN2B* | 6.50E-03 | 1.00 |
| Maturity onset diabetes of the young (26) | 2 (3.1%) | *RFX6, HNF1B* | 2.50E-03 | 1.00 |
| Huntington's disease (192) | 3 (4.6%) | *DNAH11, TP53, GRIN2B* | 0.017 | 1.00 |

## REFERENCES

1. Roehrborn CG. Pathology of benign prostatic hyperplasia. Int J Impot Res 20 Suppl 3), S11–8 (2008).

2. McNeal JE, Redwine EA, Freiha FS, Stamey TA. Zonal distribution of prostatic adenocarcinoma. Correlation with histologic pattern and direction of spread. Am J Surg Pathol 12 (12), 897–906 (1988).

3. Berry SJ, Coffey DS, Walsh PC, Ewing LL. The development of human benign prostatic hyperplasia with age. J Urol 132 (3), 474–9 (1984).

4. Siegel RL, Miller KD, Fuchs HE, Jemal A. Cancer Statistics, 2021. CA Cancer J Clin 71 (1), 7–33 (2021).

5. Alcaraz A, Hammerer P, Tubaro A, Schroder FH, Castro R. Is there evidence of a relationship between benign prostatic hyperplasia and prostate cancer? Findings of a literature review. Eur Urol 55 (4), 864–73 (2009).

6. Orsted DD, Bojesen SE. The link between benign prostatic hyperplasia and prostate cancer. Nat Rev Urol 10 (1), 49–54 (2013).

7. Sommers SC. Endocrine changes with prostatic carcinoma. Cancer 10 (2), 345–58 (1957).

8. Bostwick DG, Cooner WH, Denis L, Jones GW, Scardino PT, Murphy GP. The association of benign prostatic hyperplasia and cancer of the prostate. Cancer 70 (1 Suppl), 291–301 (1992).

9. Greenwald P, Kirmss V, Polan AK, Dick VS. Cancer of the prostate among men with benign prostatic hyperplasia. J Natl Cancer Inst 53 (2), 335–40 (1974).

10. Armenian HK, Lilienfeld AM, Diamond EL, Bross ID. Relation between benign prostatic hyperplasia and cancer of the prostate. A prospective and retrospective study. Lancet 2 (7873), 115–7 (1974).

11. Thompson IM, Coltman CA, Jr., Crowley J. Chemoprevention of prostate cancer: the Prostate Cancer Prevention Trial. Prostate 33 (3), 217–21 (1997).

12. Andriole GL, Bostwick DG, Brawley OW, Gomella LG, Marberger M, Montorsi F, et al. Effect of dutasteride on the risk of prostate cancer. N Engl J Med 362 (13), 1192–202 (2010).

13. Chokkalingam AP, Nyren O, Johansson JE, Gridley G, McLaughlin JK, Adami HO, et al. Prostate carcinoma risk subsequent to diagnosis of benign prostatic hyperplasia: a population-based cohort study in Sweden. Cancer 98 (8), 1727–34 (2003).

14. Orsted DD, Bojesen SE, Nielsen SF, Nordestgaard BG. Association of clinical benign prostate hyperplasia with prostate cancer incidence and mortality revisited: a nationwide cohort study of 3,009,258 men. Eur Urol 60 (4), 691–8 (2011).

15. Zhang L, Wang Y, Qin Z, Gao X, Xing Q, Li R, et al. Correlation between Prostatitis, Benign Prostatic Hyperplasia and Prostate Cancer: A systematic review and Meta-analysis. J Cancer 11 (1), 177–89 (2020).

16. National Cancer Institute. Understanding Prostate Changes: A Health Guide for Men 2021. https://www.cancer.gov/types/prostate/understanding-prostate-changes (accessed 3 Nov 2021)

17. Meigs JB, Barry MJ, Giovannucci E, Rimm EB, Stampfer MJ, Kawachi I. High rates of prostate-specific antigen testing in men with evidence of benign prostatic hyperplasia. Am J Med 104 (6), 517–25 (1998).

18. Lerner LB, McVary KT, Barry MJ, Bixler BR, Dahm P, Das AK, et al. Management of Lower Urinary Tract Symptoms Attributed to Benign Prostatic Hyperplasia: AUA GUIDELINE PART I-Initial Work-up and Medical Management. J Urol 206 (4), 806–17 (2021).

19. Carroll PR, Parsons JK, Andriole G, Bahnson RR, Castle EP, Catalona WJ, et al. NCCN Guidelines Insights: Prostate Cancer Early Detection, Version 2.2016. J Natl Compr Canc Netw 14 (5), 509–19 (2016).

20. Ni G, Moser G, Schizophrenia Working Group of the Psychiatric Genomics C, Wray NR, Lee SH. Estimation of Genetic Correlation via Linkage Disequilibrium Score Regression and Genomic Restricted Maximum Likelihood. Am J Hum Genet 102 (6), 1185–94 (2018).

21. Conti DV, Darst BF, Moss LC, Saunders EJ, Sheng X, Chou A, et al. Trans-ancestry genome-wide association meta-analysis of prostate cancer identifies new susceptibility loci and informs genetic risk prediction. Nat Genet 53 (1), 65–75 (2021).

22. Gudmundsson J, Sigurdsson JK, Stefansdottir L, Agnarsson BA, Isaksson HJ, Stefansson OA, et al. Genome-wide associations for benign prostatic hyperplasia reveal a genetic correlation with serum levels of PSA. Nat Commun 9 (1), 4568 (2018).

23. Zheng SL, Sun J, Wiklund F, Smith S, Stattin P, Li G, et al. Cumulative association of five genetic variants with prostate cancer. N Engl J Med 358 (9), 910–9 (2008).

24. Shi Z, Platz EA, Wei J, Na R, Fantus RJ, Wang CH, et al. Performance of Three Inherited Risk Measures for Predicting Prostate Cancer Incidence and Mortality: A Population-based Prospective Analysis. Eur Urol), (2020).

25. Bycroft C, Freeman C, Petkova D, Band G, Elliott LT, Sharp K, et al. The UK Biobank resource with deep phenotyping and genomic data. Nature 562 (7726), 203–9 (2018).

26. Yu H, Shi Z, Wu Y, Wang CH, Lin X, Perschon C, et al. Concept and benchmarks for assessing narrow-sense validity of genetic risk score values. Prostate 79 (10), 1099–105 (2019).

27. Burgess S, Butterworth A, Thompson SG. Mendelian randomization analysis with multiple genetic variants using summarized data. Genet Epidemiol 37 (7), 658–65 (2013).

28. Burgess S, Davey Smith G, Davies NM, Dudbridge F, Gill D, Glymour MM, et al. Guidelines for performing Mendelian randomization investigations. Wellcome Open Res 4), 186 (2019).

29. Xu J, Isaacs SD, Sun J, Li G, Wiley KE, Zhu Y, et al. Association of prostate cancer risk variants with clinicopathologic characteristics of the disease. Clin Cancer Res 14 (18), 5819–24 (2008).

30. Yamashiro JR, de Riese WTW. Any Correlation Between Prostate Volume and Incidence of Prostate Cancer: A Review of Reported Data for the Last Thirty Years. Res Rep Urol 13), 749–57 (2021).

